# Sex-specific socioeconomic inequalities in trajectories of anthropometry, blood pressure and blood-based biomarkers from birth to 18 years: a prospective cohort study

**DOI:** 10.1101/2023.09.27.23296220

**Authors:** Kate N O’Neill, Minhal Ahmed, Linda M O’Keeffe

## Abstract

**Background:** Evidence on when socioeconomic inequalities in conventional cardiometabolic risk factors emerge and how these change over time is sparse but important in identifying pathways leading to socioeconomic inequalities in cardiovascular disease (CVD). We examine socioeconomic inequalities in trajectories of cardiometabolic risk factors across childhood and adolescence.

**Methods:** Data were from offspring of the Avon Longitudinal Study of Parents and Children (ALSPAC), born in 1991/1992. Socioeconomic position (SEP) was measured using maternal education from questionnaires administered to mothers at 32-weeks’ gestation. Cardiometabolic risk factors were measured from birth/mid-childhood to age 18 years (y) and included fat and lean mass (9y–18y), systolic and diastolic blood pressure (SBP, DBP), pulse rate and glucose (7y-18y), high-density lipoprotein cholesterol (HDL-c), non-HDL-c and triglycerides (birth-18y). We examined the sex-specific associations between SEP and trajectories of risk factors using linear spline multilevel models.

**Results:** Among 6,517-8,952 participants with 11,948-42,607 repeated measures, socioeconomic inequalities in fat mass were evident at age 9y and persisted throughout adolescence, with graded associations across levels of SEP among females only. By 18y, fat mass was 12.32% (95% Confidence Interval (CI):6.96,17.68) lower among females and 7.94% (95% CI:1.91,13.97) lower among males with the highest SEP compared to the lowest. Socioeconomic inequalities in SBP and DBP trajectories were evident at 7y, narrowed in early adolescence and re-emerged between ages 16y-18y, particularly among females. Socioeconomic inequalities in lipid trajectories emerged, among females only, between birth and 9y in non-HDL-c, 7y and 18y in HDL-c and 9y and 18y in triglycerides while inequalities in glucose emerged among males only between ages 15y-18y.

**Conclusion:** Prevention targeting the early life course may be beneficial for reducing socioeconomic inequalities in CVD especially among females who have greater socioeconomic inequalities in cardiometabolic risk factors than males at the end of adolescence.

## Introduction

Cardiovascular disease (CVD) is the leading cause of death globally (1). CVD risk develops early in the life course (2–4) and tracks from childhood through to adulthood (5–7). Socioeconomic inequalities in CVD are well established (8–14). However, despite decades of research their prevention remains a global health challenge (15) and continues to be prioritised in national and international health strategies (12). Recent evidence has also suggested that socioeconomic inequalities are likely to widen further in the post Covid-era, thereby necessitating renewed efforts to tackle socioeconomic inequalities in CVD risk across the life course (16,17).

Efforts to prevent or reduce socioeconomic inequalities in CVD have been curbed by a limited understanding of the pathways that likely drive such inequalities across the life course, including in particular when socioeconomic inequalities in key risk factors emerge and how they change over time (18). For instance, numerous studies have examined socioeconomic inequalities in adiposity in early life (19–24), but these have often only had measures available at a single time point in childhood or have studied trajectories of adiposity from early childhood onwards but have not included risk factors such as blood pressure and lipids to examine the concomitant change together to better understand aetiology. For example, an analysis of the Avon Longitudinal Study of Parents and Children (ALSPAC) birth cohort study explored trajectories of ponderal index from birth to 2 years (y) and body mass index from 2y to 10y demonstrating that socioeconomic inequalities in adiposity emerged around the age of 4y and continued to widen with increasing age up to 10y (23). A recent analysis of the Millenium Cohort Study examined trajectories of fat mass from age 7y to 17y and found socioeconomic inequalities were evident at age 7y and continued to widen with age up to 17y (25). A further analysis of the ALSPAC cohort explored socioeconomic inequalities in trajectories of fat mass, blood pressure and lipids and found inequalities in fat mass among females only and in cholesterol and blood pressure in both sexes. However, measurements were only available from age 9y to 15y and it is not clear whether these patterns track into early adulthood (22).

Understanding the socioeconomic patterning of cardiometabolic risk factors across the life course can provide important insights into when socioeconomic inequalities emerge and how they change over time (18). Such insights may inform opportunities for the prevention of socioeconomic inequalities in CVD. We use the Avon Longitudinal Study of Children and Parents (ASLPAC), a contemporary prospective birth cohort study in the Southwest of England, to examine socioeconomic inequalities in trajectories of cardiometabolic risk factors across childhood and adolescence, from birth to 18y. Risk factors include height-adjusted fat and lean mass (9y to 18y), systolic blood pressure (SBP), diastolic blood pressure (DBP), pulse rate and glucose (7y to 18y), high density lipoprotein cholesterol (HDL-c), non-high density lipoprotein cholesterol (non-HDL-c), and triglycerides (birth to 18y).

## Methods

### Study participants

Data were from first-generation children of the Avon Longitudinal Study of Parents and Children (ALSPAC), a population-based prospective birth cohort study in southwest England (26,27). Pregnant women resident in one of three Bristol-based health districts with an expected delivery date between April 1, 1991, and December 31, 1992, were invited to participate. ALSPAC initially enrolled a cohort of 14,451 pregnancies, from which 14,062 live births occurred, and 13,988 children were alive at age 1y. When the oldest children were approximately 7y, an attempt was made to bolster the sample with eligible cases who had not joined the study originally. Therefore, the total sample size for analyses using data collected after the age of 7y is 14,901 children. Follow-up has included parent- and child-completed questionnaires, research clinic attendance, and links to routine data. Ethical approval for the study was obtained from ALSPAC Ethics and Law Committee and the Local Research Ethics Committees. Informed consent for the use of data collected via questionnaires and clinics was obtained from participants following the recommendations of the ALSPAC Ethics and Law Committee at the time. The study website contains details of all the data that is available through a fully searchable data dictionary and variable search tool - http://www.bristol.ac.uk/alspac/researchers/our-data/.

### Socioeconomic position

Maternal educational attainment was used as an indicator of socioeconomic position (SEP) as it is the most complete measure available and is the most frequently reported indicator of childhood SEP (23,24,28). At 32-weeks’ gestation, mothers were asked to report their highest educational attainment based on UK standards at the time which was categorised as ‘less than O-level’ (Ordinary Level; exams taken in different subjects usually at age 15y or 16y at the completion of legally required school attendance, equivalent to the present UK General Certificate of Secondary Education; assumed to reflect the lowest SEP), ‘O-level’, ‘A-level’ (Advanced Level; exams taken in different subjects usually at age 18y), or ‘university degree’ (undergraduate or postgraduate; assumed to reflect the highest SEP).

### Cardiometabolic risk factor measurement

#### Fat and Lean Mass

Whole body less head, and central fat and lean mass were derived from whole body dual energy X-ray absorptiometry (DXA) scans assessed five times at ages 9, 11, 13, 15, and 18y using a Lunar prodigy narrow fan beam densitometer.

#### SBP, DBP and Pulse

At each clinic (ages 7, 9, 10, 11, 12, 15 and 18y), SBP, DBP and pulse rate were measured at least twice in each with the child sitting and at rest with the arm supported, using a cuff size appropriate for the child’s upper arm circumference and a validated blood pressure monitor. The mean of the two final measures was used.

#### Blood Based Biomarkers

Non-fasting glucose was measured at age 7y as part of metabolic trait profiling, using Nuclear Magnetic Resonance (NMR) spectroscopy. In a random 10% of the cohort at age 9y, fasting glucose was also available; taken as part of a continuation of an earlier sub-study called “Child in Focus” which included approximately 10% of the overall cohort (26). Fasting glucose was available from research clinics held when participants were aged 15y and 18y. HDL-c, total cholesterol and triglycerides were measured in cord blood at birth and from venous blood subsequently. Samples were non-fasted at 7y and 9y; fasting measures were available from clinics at 15y and 18y. Non-HDL-c was calculated by subtracting HDL-c from total cholesterol at each measurement occasion. Trajectories of glucose, triglycerides, HDL-c, non-HDL were derived from a combination of measures from cord blood, fasting bloods and non-fasting bloods, with most measures obtained through standard clinical chemistry assays, but one measure (glucose at age 7y) by NMR spectroscopy.

### Statistical analysis

We used multilevel models to examine the association between SEP and change in each risk factor across childhood and adolescence. Multilevel models estimate mean trajectories of the risk factor while accounting for the non-independence (i.e. clustering) of repeated measurements within individuals, change in scale and variance of measures over time, and differences in the number and timing of measurements between individuals (29,30). Sex-specific trajectories of all outcomes have been modelled previously using multilevel models and are described elsewhere in detail (2,31–33). Briefly, trajectories of risk factors were estimated using linear spline multilevel models (all models had two levels: measurement occasion and individual). We included all participants with data on sex, SEP and at least one measurement of the risk factor throughout the study period in each multilevel model, under a missing-at-random (MAR) assumption, to minimise selection bias (30,34). Model fit statistics for each risk factor trajectory in our sample are shown in Supplementary Material Tables S1-S7.

To explore the sex-specific associations between SEP and cardiometabolic risk factors, maternal education of less than O-level was selected as the reference category in analyses. An interaction term between each of the other maternal education categories (O-levels, A-levels and university degree) and the intercept and each linear spline period was included in all models to estimate the differences in intercepts and slopes between each maternal education category and the reference group. All models included interaction terms between sex and the intercept and linear splines to allow trajectories to differ in females and males. To explore the association between SEP and cardiometabolic risk factors in females and males separately, further interaction terms were included between sex, SEP and the intercept and linear splines.

In all models, age (in years) was centred at the first available measure. Fat mass and lean mass were adjusted for height using the time- and sex-varying power of height that best resulted in a height-invariant measure, as described previously (31). Values of cardiometabolic risk factors that had a skewed distribution (fat mass, triglycerides) were (natural) log transformed prior to analyses. Differences and confidence intervals were calculated on the log-scale and were back-transformed and are therefore interpreted as the ratio of geometric means. Graphs displayed for these risk factors are in original units and derived by back transforming from the natural log scale. All trajectories were modelled in MLwiN version 3.04, called from Stata version 17 using the runmlwin command (35).

## Results

The number of participants included in the analyses ranged from a maximum of 8,952 participants (19,938 measures) for triglycerides to a minimum of 6,517 participants (11,948 measures) for glucose (Table S8, Figure S1). Characteristics of participants by sex are detailed in Table 1. Maternal education was similar in females and males, with approximately a quarter of mothers having less than O-levels educational attainment. Approximately 14% had educational attainment up to degree level or above. Compared with participants excluded from analyses, those included had higher maternal education, higher household social class and lower maternal smoking during pregnancy (Table S9).

**Table 1.**
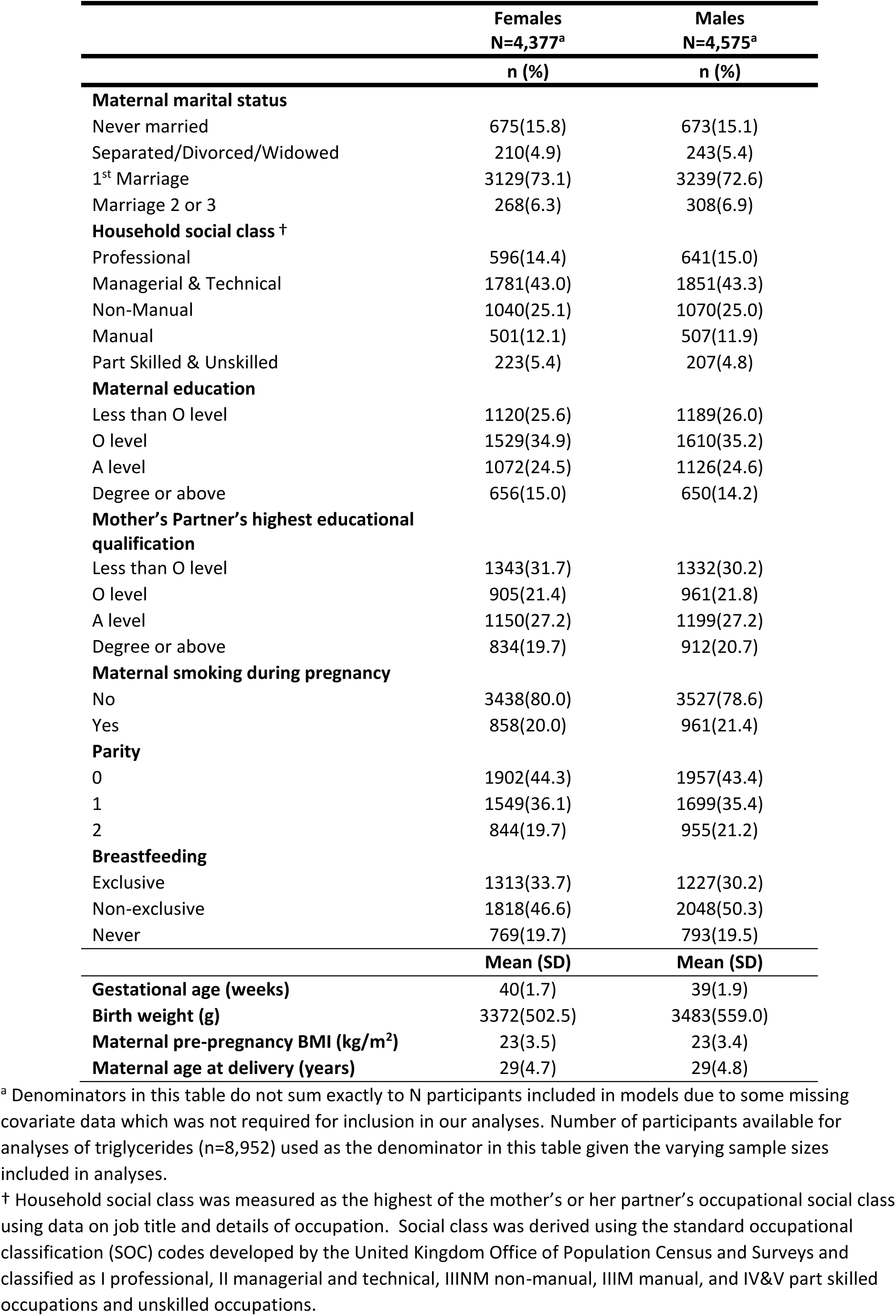
Characteristics of ALSPAC participants included in the analysis, by sex.

|  | **Females**<br>**N=4,377<sup>a</sup>** | **Males**<br>**N=4,575<sup>a</sup>** |
| --- | --- | --- |
|  | **n (%)** | **n (%)** |
| **Maternal marital status** |  |  |
| Never married | 675(15.8) | 673(15.1) |
| Separated/Divorced/Widowed | 210(4.9) | 243(5.4) |
| 1<sup>st</sup> Marriage | 3129(73.1) | 3239(72.6) |
| Marriage 2 or 3 | 268(6.3) | 308(6.9) |
| **Household social class <sup>†</sup>** |  |  |
| Professional | 596(14.4) | 641(15.0) |
| Managerial & Technical | 1781(43.0) | 1851(43.3) |
| Non-Manual | 1040(25.1) | 1070(25.0) |
| Manual | 501(12.1) | 507(11.9) |
| Part Skilled & Unskilled | 223(5.4) | 207(4.8) |
| **Maternal education** |  |  |
| Less than O level | 1120(25.6) | 1189(26.0) |
| O level | 1529(34.9) | 1610(35.2) |
| A level | 1072(24.5) | 1126(24.6) |
| Degree or above | 656(15.0) | 650(14.2) |
| **Mother's Partner's highest educational qualification** |  |  |
| Less than O level | 1343(31.7) | 1332(30.2) |
| O level | 905(21.4) | 961(21.8) |
| A level | 1150(27.2) | 1199(27.2) |
| Degree or above | 834(19.7) | 912(20.7) |
| **Maternal smoking during pregnancy** |  |  |
| No | 3438(80.0) | 3527(78.6) |
| Yes | 858(20.0) | 961(21.4) |
| **Parity** |  |  |
| 0 | 1902(44.3) | 1957(43.4) |
| 1 | 1549(36.1) | 1699(35.4) |
| 2 | 844(19.7) | 955(21.2) |
| **Breastfeeding** |  |  |
| Exclusive | 1313(33.7) | 1227(30.2) |
| Non-exclusive | 1818(46.6) | 2048(50.3) |
| Never | 769(19.7) | 793(19.5) |
|  | **Mean (SD)** | **Mean (SD)** |
| **Gestational age (weeks)** | 40(1.7) | 39(1.9) |
| **Birth weight (g)** | 3372(502.5) | 3483(559.0) |
| **Maternal pre-pregnancy BMI (kg/m<sup>2</sup>)** | 23(3.5) | 23(3.4) |
| **Maternal age at delivery (years)** | 29(4.7) | 29(4.8) |

<sup>a</sup> Denominators in this table do not sum exactly to N participants included in models due to some missing covariate data which was not required for inclusion in our analyses. Number of participants available for analyses of triglycerides (n=8,952) used as the denominator in this table given the varying sample sizes included in analyses.
<sup>†</sup> Household social class was measured as the highest of the mother's or her partner's occupational social class using data on job title and details of occupation. Social class was derived using the standard occupational classification (SOC) codes developed by the United Kingdom Office of Population Census and Surveys and classified as I professional, II managerial and technical, IIINM non-manual, IIIM manual, and IV&V part skilled occupations and unskilled occupations.

### Fat and Lean Mass

Mean fat and lean mass trajectories by maternal education are presented in Figure 1 and Table S10-S11. Among females, there was evidence of a socioeconomic gradient in fat mass across all maternal education categories at age 9y with some evidence of inequalities widening over time. For instance, at 9y fat mass was 3.0% (95% confidence interval (CI): −1.60,7.60), 4.99% (95% CI: 0.20,9.78) and 10.61% (95% CI: 5.38,15.75) lower among females with O-level, A-level and degree level maternal education, compared to females with less than O-level, respectively. By 18y, associations strengthened with 4.68% (95% CI: −0.21,9.57), 8.43% (95% CI: 3.44,13.42) and 12.32% (95% CI: 6.96,17.68) lower fat mass among females with O-level, A-level and degree level education, compared to females with less than O-level, respectively. Among males, lower fat mass was only evident in those with degree level maternal education compared with less than O-level at age 9y and associations were similar at 18y. For instance, degree level maternal education was associated with 7.08% (95% CI: −1.66,12.50) lower fat mass at age 9y and 7.94% (95% CI: 1.91,13.97) lower fat mass at 18y.

**Figure 1:**
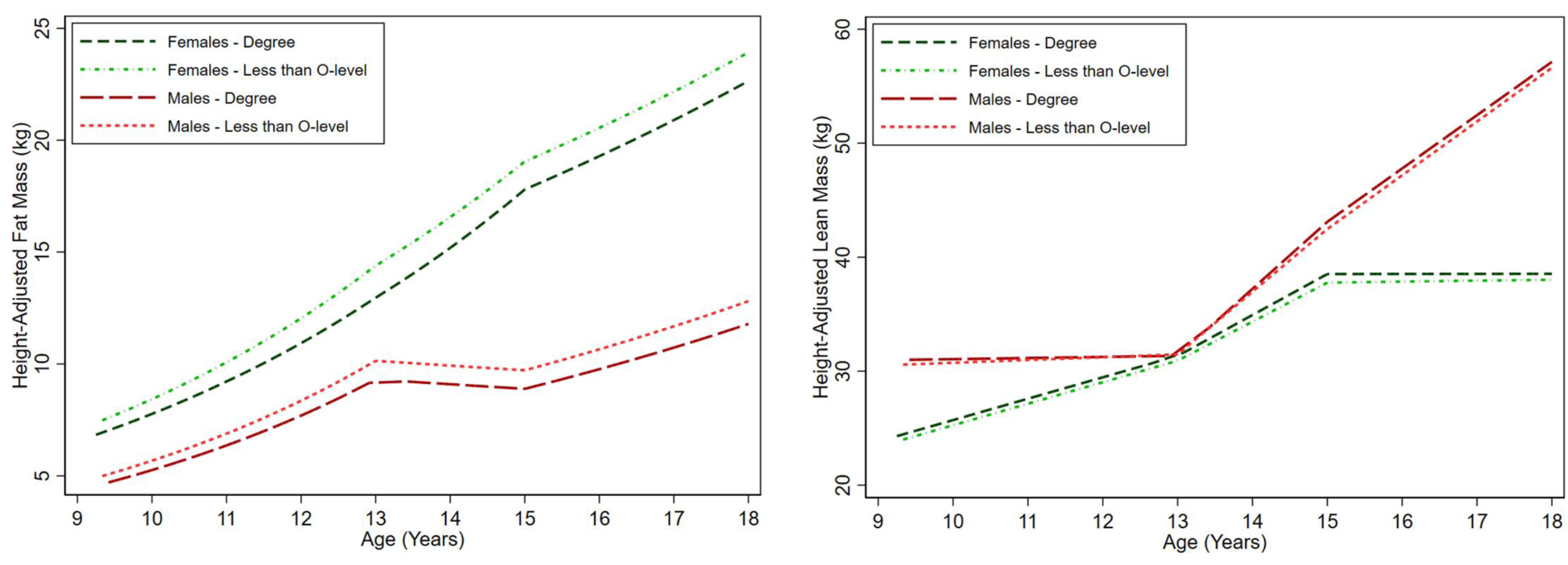
Mean trajectories of fat mass and lean mass in females and males, by maternal education, estimated from multilevel models. Detailed results with confidence intervals are provided in Supplementary Table S10-11. kg, kilograms

Among females, higher lean mass was evident among those with degree level maternal education compared to less than O-level maternal education at 18y only (difference: 0.51kg (95% CI:-0.09, 1.11)). Higher lean mass at 9y was evident among males with higher maternal education, and this persisted at 18y. For instance, at 18y, a higher lean mass of 0.87kg (95% CI: 0.29, 1.46) and 0.52kg (95% CI: −0.13,1.17) was evident among males with degree level maternal education and A-level education, respectively, compared to less than O-level.

### SBP, DBP, Pulse

Mean SBP, DBP and pulse trajectories by maternal education are presented in Figure 2 and Table S12. Among females, there was evidence of a socioeconomic gradient in SBP across all maternal education categories at age 7y. For instance, O-level, A-level and degree level maternal education were associated with 0.98mmHg (95% CI: 0.16,1.79), 1.71mmHg (95% CI: 0.85,2.57) and 2.32mmHg (95% CI: 1.34-3.29) lower SBP, respectively, compared with less than O-level. Faster increases in SBP were observed among those with degree level maternal education compared to less than O-level from 12y to 16y but from 16y to 18y there was evidence of faster decreases in SBP among O-level, A-level and degree educated categories compared with the reference group giving rise to widening inequalities by 18y. At 18y, O-level, A-level and degree level maternal education were associated with 1.30mmHg (95% CI: 0.31,2.29), 2.74mmHg (95% CI: 1.69,3.80) and 3.04mmHg (95% CI: 1.88-4.21) lower SBP, respectively among females. At age 7y, associations with were broadly similar among males with evidence of a socioeconomic gradient in SBP across all maternal education categories. For instance, O-level, A-level and degree level maternal education were associated with 0.64mmHg (95% CI: −0.16,1.43), 1.30mmHg (95% CI: 0.45,2.15) and 2.29mmHg (95% CI: 1.31-3.27) lower SBP, respectively, compared with less than O-level. Faster increases in SBP were observed among those with A-level and degree level maternal education compared to less than O-level from 7y to 12y. Faster decreases in SBP were observed among those with degree level maternal education only from 16y to 18y such that by 18y, associations had weakened across maternal education categories and lower SBP was only observed in those with degree level maternal education compared with less than O-level (difference: −1.50mmHg (95%CI:-2.80,-0.19)). Similar patterns of associations were observed for DBP.

**Figure 2:**
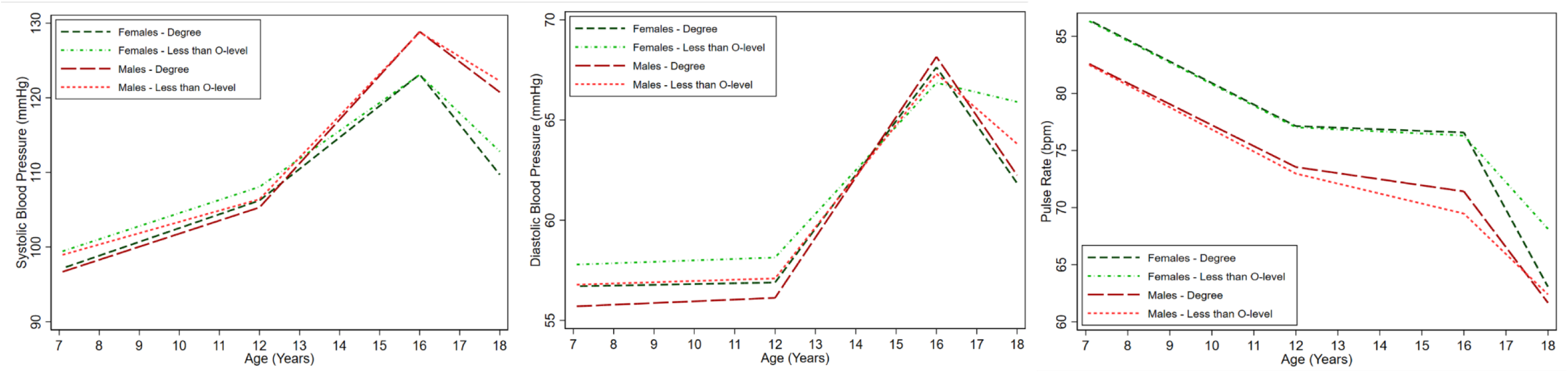
Mean trajectories of SBP, DBP and pulse in females and males, by maternal education, estimated from multilevel models. Detailed results with confidence intervals are provided in Supplementary Table S12. DBP, diastolic blood pressure; SBP, systolic blood pressure; mmHg, millimetres of mercury; bpm, beats per minute

There was no evidence of associations of maternal education and pulse at age 7y among females. Faster decreases were observed among females with degree level and A-level maternal education between ages 16y and 18y compared to those with less than O-level education resulting in a lower pulse rate of 2.74 bpm (95% CI: 1.45-4.03) and 2.74 bpm (95% CI: 1.58,3.91) respectively by age 18y. There was no evidence of socioeconomic inequalities in pulse rate trajectories from 7y to 18y among males.

### Blood Based Biomarkers

Mean glucose trajectories by maternal education are presented in Figure 3 and Table S13. There was little evidence of associations of maternal education and glucose trajectories except for 0.09mmol/L (95% CI: 0.02-0.16) lower glucose among males with degree level maternal education at age 18y compared with those with less than O-level maternal education. This was driven by faster decreases among those with degree level maternal education compared to less than O-level maternal education between ages 15y and 18y.

**Figure 3:**
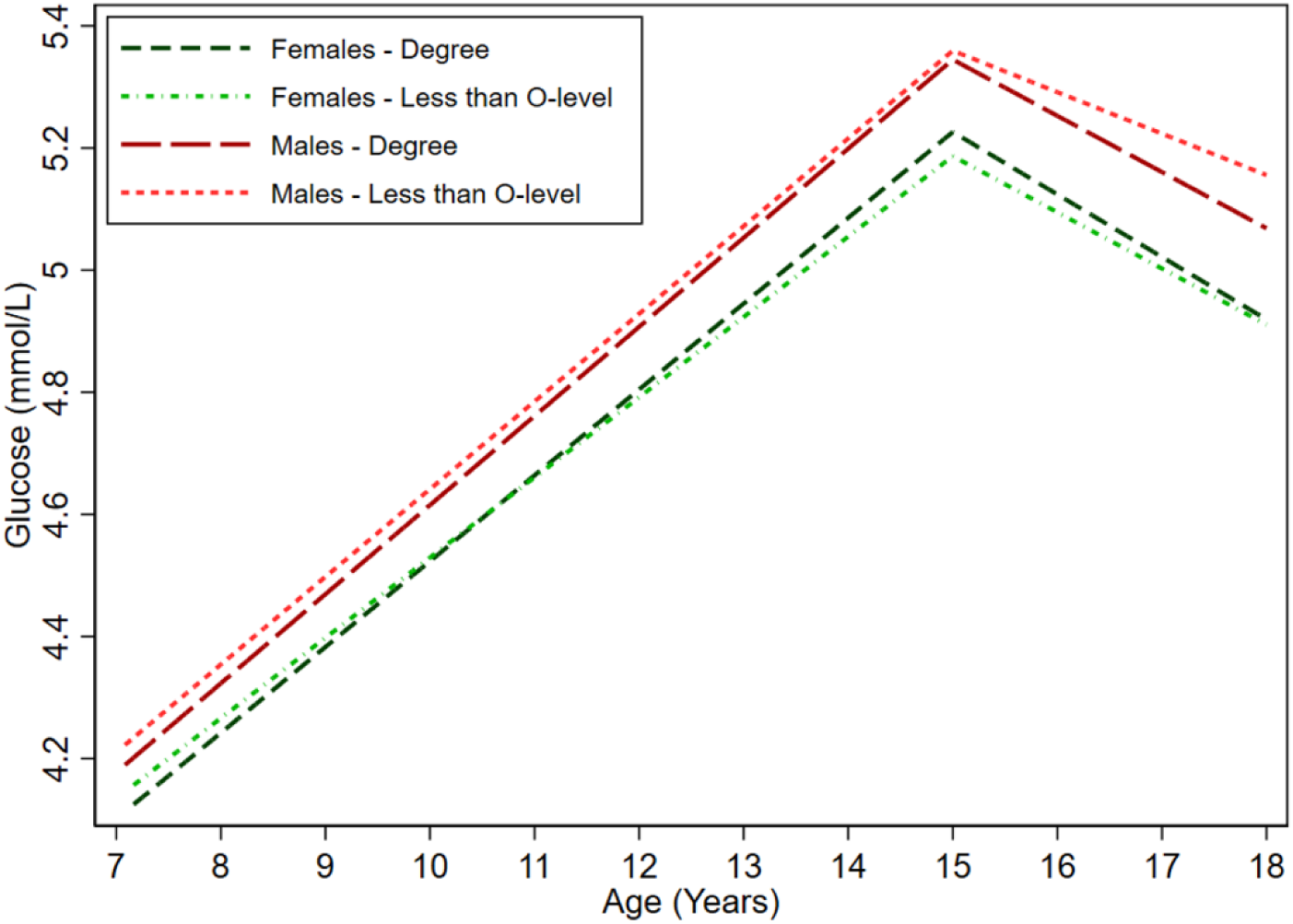
Mean trajectories of glucose in females and males, by maternal education, estimated from multilevel models. Detailed results with confidence intervals are provided in Supplementary Table S13. mmol/l, millimole per litre

Mean HDL-c, non-HDL-c and triglyceride trajectories by maternal education are presented in Figure 4 and Table S14-S15. At birth, there was little evidence of associations of maternal education and non-HDL-c trajectories among females (Figure 4, Table S14). Between 0-9y, slower increases in non-HDL-c were observed among females with higher maternal education such that by age 18y, non-HDL-c was 0.05mmol/L (95% CI:-0.02, 0.12), 0.09mmol/L (95% CI:0.01, 0.16) and 0.09 mmol/L (95% CI: 0.01, 0.18) lower among those with A-level, O-level and degree level maternal education compared with less than O-level, respectively. Maternal education was not associated with non-HDL-c trajectories from birth to 18y among males.

**Figure 4:**
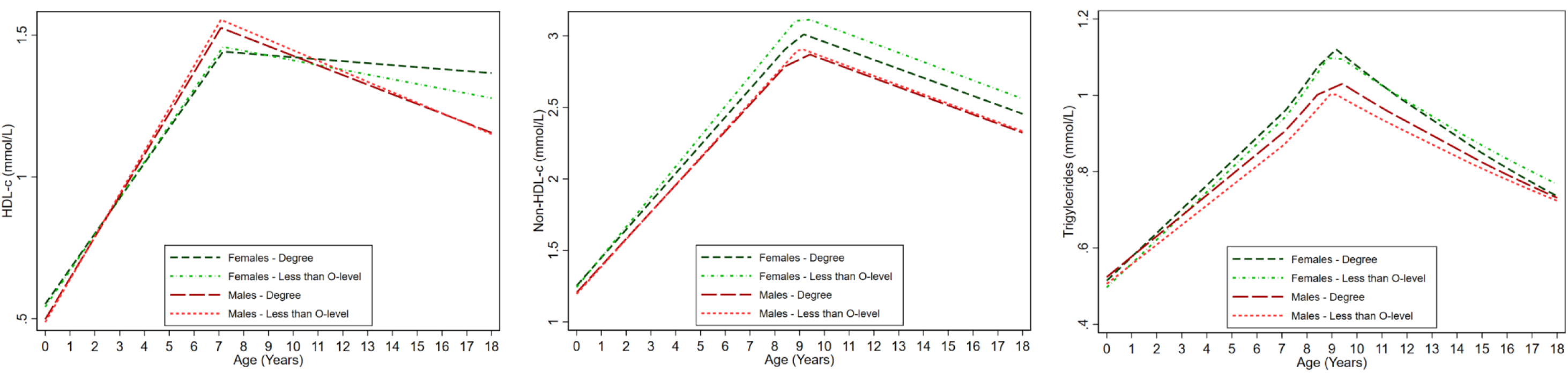
Mean trajectories of HDL-c, non-HDL-c and triglycerides in females and males, by maternal education, estimated from multilevel models. Detailed results with confidence intervals are provided in Supplementary Table S14-15. HDL-c, high density lipoprotein cholesterol; Non-HDL-c, non-high density lipoprotein cholesterol; mmol/l, millimole per litre

At birth, there was little evidence of associations of maternal education and HDL-c trajectories in both sexes (Figure 4, Table S14). Associations of maternal education and HDL-c emerged among females due to slower decreases among those with higher maternal education between 7y and 18y such that by 18y, O-level, A-level and degree level maternal education was associated with 0.03 mmol/L (95% CI: 0.001,0.06), 0.05 mmol/L (95% CI: 0.02,0.08) and 0.11 mmol/L (95% CI: 0.07,0.15) higher HDL-c compared with less than O-level, respectively. Maternal education was not associated with HDL-c trajectories from birth to 18y among males.

Similar patterns of associations were observed for triglycerides with little evidence of associations at birth (Figure 4, Table S15). However, associations of maternal education and triglycerides emerged between 9y and 18y among females only due to faster decreases among those with degree level maternal education compared with less than O-level. By 18y, degree level maternal education was associated with 7.11% (95% CI:2.35,11.86) lower triglycerides compared with less than O-level. There was little evidence of maternal education associations with triglycerides from birth to 18y among males.

## Discussion

In this large prospective birth cohort study, we examined socioeconomic inequalities in trajectories of nine cardiometabolic risk factors during childhood and adolescence. Socioeconomic inequalities in fat mass and blood pressure were evident in childhood and persisted throughout adolescence, with graded associations across all levels of maternal education among females but not males. Adolescence was an important period for the emergence of socioeconomic inequalities in pulse, HDL-c and triglycerides among females only while inequalities in glucose emerged among males only.

Similar to previous findings in ALSPAC (23,24), socioeconomic inequalities in fat mass were evident at age 9y in both females and males and there was evidence of a SEP gradient in associations among females only whereby fat mass was lower across increasing categories of maternal education. In contrast, among males, socioeconomic inequalities in fat mass were evident only in those with degree level maternal education and there was little SEP gradient across all lower levels of maternal education. We show that this childhood pattern continues into adolescence and that there was some evidence of inequalities widening across all categories of maternal education among females by 18y while remaining similar over time among males. Our findings are consistent with a recent systematic review, including 50 studies mostly from high-income countries, that reported socioeconomic inequalities in fat mass in childhood that were wider among females compared with males (19). Analyses of the Millennium Cohort Study in the UK also found greater evidence of inequalities in fat mass in females early in childhood, but in contrast to our findings, inequalities widened at a faster rate in males so that by late adolescence, inequalities in females and males were similar (25). However, area-level deprivation was used as the indicator of SEP in Millennium Cohort Study which reportedly captures elements of the environment beyond family SEP in adolescence, but not in early childhood. Taken together, these findings may suggest that different dimensions of SEP influence risk in females and males across the early life course. SEP at different points throughout the life course has previously been shown to influence CVD risk differently in adult females and males (36). We found little evidence of socioeconomic inequalities in height-adjusted lean mass, consistent with previous literature (19).

Similar to patterns of socioeconomic inequalities in fat mass and a previous ALSPAC analysis that explored associations of maternal education and blood pressure at age 10y (24), we found socioeconomic inequalities in blood pressure in childhood. A further analysis of approximately 8,500 children in ALSPAC demonstrated that inequalities in blood pressure narrowed with age from 7y to 15y (22). However, while our findings also suggest that inequalities in blood pressure narrowed throughout childhood and early adolescence, our results extend and build on the analyses of this previous study. We found faster rates of decreases in SBP and DBP among those with higher maternal education between the ages of 16y and 18y that resulted in a re-emergence of socioeconomic inequalities in SBP and DBP by age 18y, particularly among females. Similar to patterns observed in fat mass, associations were graded across levels of maternal education among females but not males. It has been posited that early influences of SEP on child health may be reduced in adolescence due to the influence of school environment, peer support and youth culture, in what has been termed an equalisation of health during youth, but that inequalities re-emerge thereafter (37). However, further work is required to determine the underlying mechanisms and whether changes over time are indeed explained by a theory of equalisation or by biological phenomenon such as the transient effect of puberty on blood pressure (38), the timing of which may differ by SEP (39). Our findings also identify sex-specific patterns in socioeconomic inequalities in lipids. Previous research in ALSPAC reported little evidence of socioeconomic inequalities in cholesterol and triglycerides at age 9y in females and males (24). However, when we analysed trajectories of HDL-c, non-HDL-c and triglycerides from birth to 18y we found that socioeconomic inequalities emerged in HDL-c and triglycerides in adolescence, and in non-HDL-c in childhood among females only. Similar to patterns observed for fat mass and blood pressure, associations were graded across categories of maternal education.

Overall, our findings are consistent with studies that demonstrate socioeconomic inequalities in CVD in adults (11,40,41), particularly those indicating that early life is an important period for the establishment of socioeconomic inequalities in CVD (36,42,43) and that inequalities in CVD are wider among females than males (40). For instance, associations of SEP with fat mass and blood pressure were graded across all levels of maternal education by 18y among females but associations were only observed for degree level maternal education compared with less than O-level among males. Furthermore, socioeconomic inequalities were observed in lipids among females only and not among males. Taken together, our findings provide important insights into the potential aetiological pathways for socioeconomic inequalities in CVD. The different patterns of socioeconomic inequalities in fat mass and blood pressure in adolescence indicate that adiposity may not fully explain the observed associations of SEP and blood pressure. Inequalities in fat mass persisted among males and somewhat strengthened among females between 9y and 18y while inequalities in SBP and DBP narrowed between the ages of 7y and 12y among females and 12y and 16y among males only to re-emerge between the ages of 16y and 18y. In contrast, the emergence of socioeconomic inequalities in lipids among females in adolescence particularly may indicate a role for adiposity given the suggestive evidence of widening inequalities in fat mass among females over this period. However, future work exploiting the maturation of contemporary birth cohort studies alongside advancements in methods for mediation analysis (44) is required to determine whether inequalities track into adulthood and explain the mediating pathways through which early life SEP influences CVD risk. Such work should also explore mechanisms of the differential patterns of associations observed over time and between the sexes, including the role of life course SEP, to improve our understanding of how socially structured exposures become embodied and to thus identify potential intervention targets at appropriate timepoints across the life course (18,45,46).

### Strengths and limitations

There are a number of strengths to our study including the availability of repeated measures of cardiometabolic risk factors from birth/early life to age 18y and the use of multilevel models allowing for clustering of repeated measures within individuals and correlation between measures over time. Multilevel models also allow for the inclusion of all participants with at least one measure of a cardiometabolic risk factor throughout the follow-up period, thereby minimising potential for selection bias and increasing our sample size. The study also has some limitations. The possibility of selection bias remains given that participants included in our analysis were more likely to have higher maternal education levels compared with those excluded due to missing data. Loss-to-follow up is another potential limitation of our study. However, we have included all participants with at least one measure of each risk factor to minimise any potential selection bias. The number of people with measurements of each risk factor also varied meaning that our sample sizes differed for each risk factor and thus are not directly comparable. ALSPAC is a contemporary prospective cohort study, however SEP data refers to childhood SEP in the early 1990s. The meaning of SEP indicators such as education change over time and as such likely cohort effects need to be considered in the interpretation of findings (47).

## Conclusion

Socioeconomic inequalities in fat mass and blood pressure emerge in childhood for both sexes and in adolescence for lipids and pulse rate among females and glucose among males, with some evidence of stronger graded associations among females compared with males. Prevention targeting the early life course may be beneficial for reducing socioeconomic inequalities in CVD especially among females who have greater socioeconomic inequalities in cardiometabolic risk factors than males at the end of adolescence.

## Data Availability

While the ALSPAC data used in these analyses are de-identified, there are legal restrictions on sharing these data imposed by the custodians of the data, The University of Bristol. Further information can be found here: http://www.bristol.ac.uk/media-library/sites/alspac/documents/researchers/data-access/ALSPAC_Access_Policy.pdf. All data enquiries can be sent to; Tel: +44 (0)117 331 0167.

## Acknowledgements

We are extremely grateful to all the families who took part in this study, the midwives for their help in recruiting them, and the whole ALSPAC team, which includes interviewers, computer and laboratory technicians, clerical workers, research scientists, volunteers, managers, receptionists and nurses.

