## Supplementary Material for "Sex-specific socioeconomic inequalities in trajectories of anthropometry, blood pressure and blood-based biomarkers from birth to 18 years: a prospective cohort study"

**Supplementary Material Contents**

**Table S1:** Model details for log fat mass trajectories
**Table S2:** Model details for lean mass trajectories
**Table S3:** Model details for SBP, DBP and pulse rate trajectories
**Table S4:** Model details for glucose trajectories
**Table S5:** Model details for HDL-c trajectories
**Table S6:** Model details for non-HDL-c trajectories
**Table S7:** Model details for log triglyceride trajectories
**Figure S1:** Flow diagram of study
**Table S8:** Number of participants with cardiometabolic risk factor measures at each time point
**Table S9:** Characteristics at birth of the mothers of children included in models compared with those excluded due to missing exposure or outcome data
**Table S10:** Mean trajectories of log fat mass and mean differences by maternal education, estimated from multilevel models
**Table S11:** Mean trajectories of lean mass and mean differences by maternal education, estimated from multilevel models
**Table S12:** Mean trajectories of SBP, DBP and pulse and mean differences by maternal education, estimated from multilevel models
**Table S13:** Mean trajectories of glucose and mean differences by maternal education, estimated from multilevel models
**Table S14:** Mean trajectories of HDL-c and non-HDL-c and mean differences by maternal education, estimated from multilevel models
**Table S15:** Mean trajectories of log triglycerides and mean differences by maternal education, estimated from multilevel models

**Table S1:** Model details for log fat mass trajectories

| | Number of contributing individuals | | Assessment of model fit | | | |
|---|---|---|---|---|---|---|
| | Total observations | Individuals with 1 measure | Mean observed, ln(kg) (SD)[a] | Mean predicted, ln(kg) (SD)[aa] | Mean difference (observed – predicted), ln(kg)[a] | 95% level of agreement between observed and predicted, ln(kg)[a] |
| Overall | 27609 | 7887 | | | | |
| 9 years | 6602 | 6602 | 1.97 (0.57) | 1.97 (0.54) | -0.002 | -0.18 to 0.17 |
| 9-13 years | 12972 | 7426 | 2.13 (0.59) | 2.13 (0.56) | -0.0004 | -0.20 to 0.20 |
| 13-15 years | 5573 | 5531 | 2.44 (0.59) | 2.44 (0.56) | 0.002 | -0.25 to 0.25 |
| 15-18 years | 9064 | 5607 | 2.64 (0.61) | 2.64 (0.58) | -0.001 | -0.18 to 0.18 |

SD, standard deviation; ln(kg), natural log of kilograms

[a]Fat mass is presented in the natural log and values represent the mean predicted natural log of fat mass.

**Table S2:** Model details for lean mass trajectories

| | Number of contributing individuals | | Assessment of model fit | | | |
|---|---|---|---|---|---|---|
| | Total observations | Individuals with 1 measure | Mean observed, kg (SD) | Mean predicted, kg (SD) | Mean difference (observed – predicted), kg | 95% level of agreement between observed and predicted, kg |
| Overall | 27676 | 7900 | | | | |
| 9 years | 6619 | 6619 | 24.55 (3.20) | 24.49 (2.72) | 0.06 | -3.30 to 3.43 |
| 9-13 years | 13001 | 7439 | 27.10 (4.62) | 27.12 (4.30) | -0.03 | -3.15 to 3.09 |
| 13-15 years | 5583 | 5541 | 37.97 (6.43) | 37.90 (6.48) | 0.07 | -3.62 to 3.75 |
| 15-18 years | 9092 | 5621 | 44.25 (9.26) | 44.25 (8.85) | -0.003 | -2.19 to 2.18 |

SD, standard deviation; kg, kilograms

**Table S3:** Model details for SBP, DBP and pulse rate trajectories

| | Number of contributing individuals | | Assessment of model fit | | | |
|---|---|---|---|---|---|---|
| | Total observations | Individuals with 1 measure | Mean observed SBP, DBP or pulse rate (SD) [a] | Mean predicted SBP, DBP or pulse rate (SD) [a] | Mean difference (observed–predicted) [a] | 95% level of agreement between observed and predicted [a] |
| **SBP** | | | | | | |
| Overall | 42368 | 8801 | | | | |
| 7 years | 7320 | 7320 | 98.83 (9.13) | 98.71 (5.38) | 0.12 | -10.63 to 10.88 |
| 7-12 years | 26674 | 8499 | 102.60 (9.53) | 102.77 (6.18) | -0.17 | -11.81 to 11.48 |
| 12-16 years | 11153 | 6573 | 115.75 (11.67) | 115.30 (8.74) | 0.45 | -11.31 to 12.21 |
| 16-18 years | 4541 | 4396 | 116.89 (10.14) | 117.21 (7.34) | -0.32 | -12.59 to 11.96 |
| **DBP** | | | | | | |
| Overall | 42370 | 8801 | | | | |
| 7 years | 7318 | 7318 | 56.45 (6.59) | 56.68 (3.40) | -0.23 | -8.96 to 8.50 |
| 7-12 years | 26688 | 8498 | 58.10 (6.97) | 56.96 (3.40) | 1.15 | -9.12 to 11.41 |
| 12-16 years | 11147 | 6573 | 61.11 (9.58) | 61.99 (5.11) | -0.87 | -12.45 to 10.71 |
| 16-18 years | 4535 | 4388 | 64.25 (6.11) | 64.57 (3.64) | -0.33 | -13.09 to 12.44 |
| **Pulse** | | | | | | |
| Overall | 42607 | 8810 | | | | |
| 7 years | 7315 | 7315 | 83.27 (10.71) | 83.26 (6.13) | 0.01 | -13.18 to 13.21 |
| 7-12 years | 26905 | 8510 | 77.63 (11.60) | 78.98 (6.99) | -1.35 | -16.07 to 13.37 |
| 12-16 years | 11163 | 6577 | 74.04 (11.25) | 74.13 (6.91) | -0.09 | -14.04 to 13.87 |
| 16-18 years | 4539 | 4393 | 65.81 (10.16) | 66.06 (6.26) | -0.25 | -14.68 to 14.18 |

DBP, diastolic blood pressure; SBP, systolic blood pressure; SD, standard deviation

[a]units are presented in mmHg for SBP and DBP and bpm for pulse rate

**Table S4:** Model details for glucose trajectories

| | Number of contributing individuals | | Assessment of model fit | | | |
|---|---|---|---|---|---|---|
| | Total observations | Individuals with 1 measure | Mean observed, mmol/l (SD) | Mean predicted, mmol/l (SD) | Mean difference (observed – predicted), mmol/l | 95% level of agreement between observed and predicted, mmol/l |
| Overall | 11948 | 6517 | | | | |
| 7 years | 4965 | 4965 | 4.18 (0.50) | 4.22 (0.23) | -0.04 | -0.60 to 0.53 |
| 7-15 years | 5822 | 5168 | 4.29 (0.55) | 4.28 (0.26) | 0.01 | -0.59 to 0.62 |
| 15-18 years | 6126 | 4123 | 5.12 (0.39) | 5.13 (0.16) | -0.01 | -0.65 to 0.63 |

SD, standard deviation; mmol/l, millimole per litre

**Table S5:** Model details for HDL-c trajectories

| | Number of contributing individuals | | Assessment of model fit | | | |
|---|---|---|---|---|---|---|
| | Total observations | Individuals with 1 measure | Mean observed, mmol/l (SD) | Mean predicted, mmol/l (SD) | Mean difference (observed – predicted), mmol/l | 95% level of agreement between observed and predicted, mmol/l |
| Overall | 19938 | 8928 | | | | |
| Birth | 4183 | 4183 | 0.52 (0.23) | 0.52 (0.10) | -0.000001 | -0.26 to 0.26 |
| 0-7 years | 4185 | 4185 | 1.06 (0.56) | 1.05 (0.52) | 0.01 | -0.25 to 0.26 |
| 7-18 years | 15753 | 7153 | 1.39 (0.32) | 1.39 (0.24) | -0.00002 | -0.25 to 0.25 |

HDL-c, high density lipoprotein cholesterol; SD, standard deviation; mmol/l, millimole per litre

**Table S6:** Model details for non-HDL-c trajectories

| | **Number of contributing individuals** | | **Assessment of model fit** | | | |
|---|---|---|---|---|---|---|
| | Total observations | Individuals with 1 measure | Mean observed, mmol/l (SD) | Mean predicted, mmol/l (SD) | Mean difference (observed – predicted), mmol/l | 95% level of agreement between observed and predicted, mmol/l |
| Overall | 19878 | 8919 | | | | |
| Birth | 4128 | 4128 | 1.22 (0.53) | 1.23 (0.18) | -0.01 | -0.72 to 0.70 |
| 0-9 years | 9045 | 7460 | 2.07 (0.98) | 2.01 (0.77) | 0.06 | -0.63 to 0.74 |
| 9-18 years | 10833 | 5977 | 2.63 (0.67) | 2.68 (0.50) | -0.05 | -0.66 to 0.56 |

Non-HDL-c, non-high density lipoprotein cholesterol; SD, standard deviation; mmol/l, millimole per litre

**Table S7:** Model details for log triglyceride trajectories

| | Number of contributing individuals | | Assessment of model fit | | | |
|---|---|---|---|---|---|---|
| | Total observations | Individuals with 1 measure | Mean observed, ln(trig), (SD)[a] | Mean predicted, ln(trig), (SD)[a] | Mean difference (observed – predicted), ln(trig)[a] | 95% level of agreement between observed and predicted, ln(trig)[a] |
| Overall | 19938 | 8952 | | | | |
| Birth | 4267 | 4267 | -0.68 (0.45) | -0.68 (0.21) | -0.003 | -0.47 to 0.46 |
| 0-9 years | 9160 | 7510 | -0.35 (0.54) | -0.35 (0.37) | 0.01 | -0.52 to 0.53 |
| 9-18 years | 10778 | 5961 | -0.14 (0.41) | -0.14 (0.22) | -0.005 | -0.54 to 0.53 |

ln(trig), natural log of triglyceride; SD, standard deviation

[a]Triglyceride is presented in the natural log and values represent the mean predicted natural log of triglyceride at each age shown.

**Table S8:** Number of participants with cardiometabolic risk factor measures at each time point

| | Birth | Age 7 | Age 9 | Age 10 | Age 11 | Age 12 | Age 13 | Age 15 | Age 18 |
|---|---|---|---|---|---|---|---|---|---|
| Fat mass | | | 6,602 | | 6,364 | | 5,539 | 4,734 | 4,370 |
| Lean mass | | | 6,619 | | 6,376 | | 5,549 | 4,750 | 4,382 |
| SBP | | 7,320 | 6,920 | 6,556 | 6,405 | 6,069 | | 4,879 | 4,219 |
| DBP | | 7,318 | 6,927 | 6,562 | 6,410 | 6,066 | | 4,874 | 4,213 |
| Pulse rate | | 7,315 | 6,922 | 6,794 | 6,402 | 6,100 | | 4,855 | 4,219 |
| Glucose | | 4,965 | 824 | | | | | 3,181 | 2,978 |
| HDL-c | 4,183 | 4,919 | 4,657 | | | | | 3,192 | 2,987 |
| Non-HDL-c | 4,128 | 4,918 | 4,654 | | | | | 3,192 | 2,986 |
| Triglycerides | 4,267 | 4,894 | 4,629 | | | | | 3,180 | 2,968 |

HDL, high density lipoprotein cholesterol; DBP, diastolic blood pressure; SBP, systolic blood pressure.

**Figure S1: Flow diagram of study**

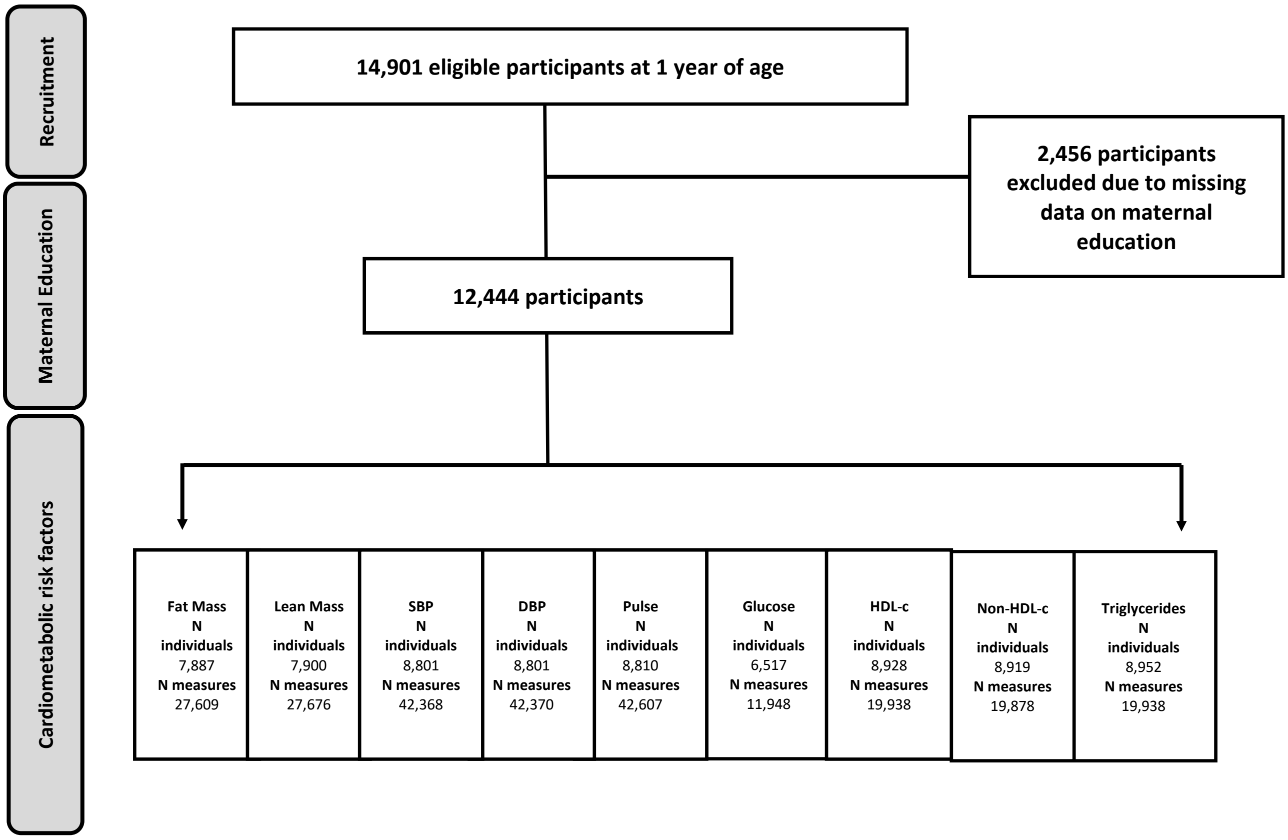


SBP, systolic blood pressure; DBP, diastolic blood pressure; HDL-c, high density lipoprotein cholesterol.

**Table S9:** Characteristics at birth of the mothers of children included in models compared with those excluded due to missing exposure or outcome data

| | Included[a] | Excluded[a] |
|---|---|---|
| | n (%) | n (%) |
| **Maternal marital status** | | |
| Never married | 1348(15.4) | 1222(25.9) |
| Separated/Divorced/Widowed | 453(5.2) | 356(7.5) |
| 1st Marriage | 6368(72.8) | 2844(60.2) |
| Marriage 2 or 3 | 576(6.6) | 302(6.4) |
| **Household social class** † | | |
| Professional | 1237(14.7) | 297(9.5) |
| Managerial & Technical | 3632(43.2) | 1179(37.8) |
| Non-Manual | 2110(25.1) | 832(26.6) |
| Manual | 1008(12.0) | 552(17.7) |
| Part Skilled & Unskilled | 430(5.1) | 262(8.4) |
| **Maternal education** | | |
| Less than O level | 2309(25.8) | 1434(41.1) |
| O level | 3139(35.1) | 1172(33.6) |
| A level | 2198(24.6) | 589(16.9) |
| Degree or above | 1306(14.5) | 298(8.5) |
| **Mother's partner's education** | | |
| Less than O level | 2675(31.0) | 1464(44.0) |
| O level | 1866(21.6) | 682(20.5) |
| A level | 2349(27.2) | 759(22.8) |
| Degree or Above | 1746(20.2) | 425(12.8) |
| **Maternal smoking during pregnancy** | | |
| No | 6965(79.3) | 2972(66.2) |
| Yes | 1891(20.7) | 1519(33.8) |
| **Parity** | | |
| 0 | 3859(43.8) | 1975(30.1) |
| 1 | 3148(35.7) | 1416(21.6) |
| 2+ | 1799(20.4) | 3175(48.4) |
| **Breastfeeding** | | |
| Exclusive | 2540(31.9) | 667(22.3) |
| Non-exclusive | 3866(48.5) | 1454(46.2) |
| Never | 1562(19.6) | 980(31.3) |
| | **Mean (SD)** | **Mean (SD)** |
| **Gestational age (weeks)** | 39(1.8) | 37(8.3) |
| **Birth weight (g)** | 3429(535.2) | 3293(643.8) |
| **Maternal pre-pregnancy BMI (kg/m$^2$)** | 23(3.5) | 23(3.6) |
| **Maternal age at delivery (years)** | 29(4.7) | 27(5.2) |

[a] Denominators for excluded participants in this table vary due to different rates of missing data for characteristics shown. Number of participants included in analyses of triglycerides (n=8,952) used as the denominator in this table given the varying sample sizes included in analyses.

† Household social class was measured as the highest of the mother's or her partner's occupational social class using data on job title and details of occupation. Social class was derived using the standard occupational classification (SOC) codes developed by the United Kingdom Office of Population Census and Surveys and classified as I professional, II managerial and technical, IIINM non-manual, IIIM manual, and IV&V part skilled occupations and unskilled occupations.

**Table S10:** Mean trajectories of log fat mass and mean differences by maternal education, estimated from multilevel models

| | Mean trajectory (95% CI) in less than O-level maternal education (reference)[a] | Mean difference in trajectory (95% CI) comparing with less than O-level maternal education[b] | | |
|---|---|---|---|---|
| | | O-level maternal education | A-level maternal education | Degree maternal education |
| **Females** | | | | |
| Age 9yr (kg) or (%) | 1.95 (1.91,1.99) | -3.00 (-7.60,1.60) | -4.99 (-9.78,-0.20) | -10.61 (-15.75,-5.48) |
| Change 9-13yr (kg/yr) or (%/yr) | 0.18 (0.17,0.19) | -0.82 (-1.88,0.25) | -1.32 (-2.43,-0.21) | -0.79 (-2.03,0.46) |
| Change 13-15yr (kg/yr) or (%/yr) | 0.14 (0.12,0.16) | 0.94 (-1.08,2.95) | 1.03 (-1.09,3.14) | 1.88 (-0.47,4.23) |
| Change 15-18yr (kg/yr) or (%/yr) | 0.08 (0.06,0.09) | -0.11 (-1.58,1.36) | -0.14 (-1.67,1.39) | -0.83 (-2.49,0.83) |
| Age 18yr (kg) or (%) | 3.17 (3.13,3.22) | -4.68 (-9.57,0.21) | -8.43 (-13.42,-3.44) | -12.32 (-17.68,-6.96) |
| **Males** | | | | |
| Age 9yr (kg) or (%) | 1.54 (1.50,1.58) | 2.09 (-2.77,6.94) | 3.41 (-1.83,8.65) | -7.08 (-12.50,-1.66) |
| Change 9-13yr (kg/yr) or (%/yr) | 0.19 (0.18,0.20) | -0.23 (-1.31,0.85) | -0.40 (-1.53,0.74) | -0.31 (-1.59,0.97) |
| Change 13-15yr (kg/yr) or (%/yr) | -0.02 (-0.04,0.00) | -0.69 (-2.74,1.36) | -1.87 (-3.97,0.23) | -0.16 (-2.54,2.22) |
| Change 15-18yr (kg/yr) or (%/yr) | 0.09 (0.08,0.11) | -0.02 (-1.69,1.66) | 0.63 (-1.08,2.33) | 0.21 (-1.62,2.05) |
| Age 18yr (kg) or (%) | 2.55 (2.50,2.60) | -0.28 (-5.91,5.35) | -0.14 (-6.01,5.72) | -7.94 (-13.97,-1.91) |

CI, confidence interval; kg/yr, kilograms per year; %/yr, percentage per year.

[a]Fat mass was transformed using the natural log. All predicted mean values (kg) and rates of change per year (kg/yr) are on the log scale

[b]The difference between maternal education categories is back transformed from the log scale for ease of interpretation and is interpreted as the percentage difference in the mean level comparing each category with less than O-level maternal education or percentage difference in change per year (%/yr) comparing each category with maternal education.

**Table S11:** Mean trajectories of lean mass and mean differences by maternal education, estimated from multilevel models

| | Mean trajectory (95% CI) in less than O-level maternal education (reference) | Mean difference in trajectory (95% CI) comparing with less than O-level maternal education | | |
|---|---|---|---|---|
| | | O-level maternal education | A-level maternal education | Degree maternal education |
| **Females** | | | | |
| Age 9yr (kg) | 23.37 (23.17,23.57) | -0.07 (-0.29,0.16) | -0.01 (-0.25,0.23) | 0.03 (-0.24,0.29) |
| Change 9-13yr (kg/yr) | 1.89 (1.79,1.98) | -0.04 (-0.14,0.06) | -0.03 (-0.14,0.07) | -0.01 (-0.13,0.11) |
| Change 13-15yr (kg/yr) | 3.43 (3.26,3.60) | 0.12 (-0.07,0.32) | 0.09 (-0.12,0.29) | 0.17 (-0.06,0.39) |
| Change 15-18yr (kg/yr) | 0.09 (-0.04,0.21) | -0.01 (-0.16,0.15) | 0.05 (-0.11,0.22) | 0.07 (-0.11,0.24) |
| Age 18yr (kg) | 38.02 (37.62,38.42) | 0.01 (-0.50,0.52) | 0.20 (-0.34,0.74) | 0.51 (-0.09,1.11) |
| **Males** | | | | |
| Age 9yr (kg) | 30.49 (30.25,30.74) | 0.18 (-0.04,0.40) | 0.33 (0.09,0.57) | 0.47 (0.19,0.74) |
| Change 9-13yr (kg/yr) | 0.25 (0.15,0.34) | -0.02 (-0.12,0.08) | -0.05 (-0.16,0.06) | -0.15 (-0.27,-0.03) |
| Change 13-15yr (kg/yr) | 5.49 (5.33,5.66) | 0.12 (-0.08,0.32) | 0.30 (0.09,0.51) | 0.38 (0.15,0.61) |
| Change 15-18yr (kg/yr) | 4.72 (4.56,4.87) | -0.04 (-0.22,0.13) | 0.05 (-0.13,0.23) | -0.04 (-0.23,0.16) |
| Age 18yr (kg) | 56.62 (56.16,57.07) | 0.22 (-0.34,0.79) | 0.87 (0.29,1.46) | 0.52 (-0.13,1.17) |

CI, confidence interval; kg/yr, kilograms per year

**Table S12:** Mean trajectories of SBP, DBP and pulse and mean differences by maternal education, estimated from multilevel models

| | Mean trajectory (95% CI) in less than O-level maternal education (reference)[a] | Mean difference in trajectory (95% CI) comparing with less than O-level maternal education | | |
|---|---|---|---|---|
| | | O-level maternal education | A-level maternal education | Degree maternal education |
| **SBP** | | | | |
| **Females** | | | | |
| Age 7yr (mmHg) | 99.29 (98.65,99.93) | -0.98 (-1.79,-0.16) | -1.71 (-2.57,-0.85) | -2.32 (-3.29,-1.34) |
| Change 7-12yr (mmHg/yr) | 1.75 (1.60,1.91) | 0.08 (-0.12,0.28) | 0.13 (-0.08,0.34) | 0.09 (-0.14,0.33) |
| Change 12-16yr (mmHg/yr) | 3.76 (3.49,4.02) | 0.13 (-0.20,0.46) | -0.03 (-0.38,0.31) | 0.47 (0.08,0.85) |
| Change 16-18yr (mmHg/yr) | -5.14 (-5.71,-4.57) | -0.63 (-1.35,0.10) | -0.78 (-1.54,-0.01) | -1.53 (-2.38,-0.68) |
| Age 18yr (mmHg) | 112.81 (112.03,113.59) | -1.30 (-2.29,-0.31) | -2.74 (-3.80,-1.69) | -3.04 (-4.21,-1.88) |
| **Males** | | | | |
| Age 7yr (mmHg) | 98.83 (98.20,99.45) | -0.64 (-1.43,0.16) | -1.30 (-2.15,-0.45) | -2.29 (-3.27,-1.31) |
| Change 7-12yr (mmHg/yr) | 1.52 (1.36,1.67) | 0.10 (-0.09,0.29) | 0.23 (0.03,0.44) | 0.24 (0.00,0.47) |
| Change 12-16yr (mmHg/yr) | 5.59 (5.31,5.87) | 0.21 (-0.14,0.56) | 0.16 (-0.20,0.52) | 0.30 (-0.11,0.70) |
| Change 16-18yr (mmHg/yr) | -3.26 (-3.92,-2.61) | -0.81 (-1.62,0.00) | -0.36 (-1.19,0.48) | -0.79 (-1.71,0.13) |
| Age 18yr (mmHg) | 122.26 (121.33,123.18) | -0.94 (-2.09,0.21) | -0.21 (-1.39,0.98) | -1.50 (-2.80,-0.19) |
| **DBP** | | | | |
| **Females** | | | | |
| Age 7yr (mmHg) | 57.78 (57.32,58.25) | -0.41 (-1.00,0.19) | -0.95 (-1.57,-0.32) | -1.36 (-2.07,-0.65) |
| Change 7-12yr (mmHg/yr) | 0.07 (-0.05,0.19) | 0.04 (-0.11,0.19) | 0.08 (-0.08,0.24) | -0.03 (-0.21,0.15) |
| Change 12-16yr (mmHg/yr) | 2.18 (1.95,2.40) | 0.19 (-0.10,0.47) | -0.01 (-0.31,0.28) | 0.51 (0.17,0.84) |
| Change 16-18yr (mmHg/yr) | -0.47 (-0.97,0.02) | -0.64 (-1.26,-0.02) | -0.60 (-1.26,0.06) | -1.28 (-2.01,-0.54) |
| Age 18yr (mmHg) | 65.91 (65.34,66.47) | -0.76 (-1.48,-0.05) | -1.83 (-2.59,-1.06) | -2.05 (-2.90,-1.21) |
| **Males** | | | | |
| Age 7yr (mmHg) | 56.78 (56.32,57.24) | -0.41 (-0.99,0.17) | -0.69 (-1.31,-0.07) | -1.08 (-1.79,-0.37) |
| Change 7-12yr (mmHg/yr) | 0.06 (-0.06,0.18) | 0.08 (-0.07,0.23) | 0.09 (-0.07,0.24) | 0.02 (-0.16,0.20) |
| Change 12-16yr (mmHg/yr) | 2.55 (2.31,2.80) | 0.38 (0.07,0.68) | 0.37 (0.06,0.69) | 0.45 (0.10,0.81) |
| Change 16-18yr (mmHg/yr) | -1.75 (-2.30,-1.19) | -0.83 (-1.52,-0.14) | -0.96 (-1.68,-0.24) | -1.21 (-2.00,-0.41) |
| Age 18yr (mmHg) | 63.81 (63.14,64.48) | -0.17 (-1.01,0.66) | -0.70 (-1.56,0.17) | -1.57 (-2.52,-0.62) |
| **Pulse rate** | | | | |
| **Females** | | | | |
| Age 7yr (bpm) | 86.49 (85.73,87.24) | -0.74 (-1.70,0.23) | -0.89 (-1.90,0.13) | -0.52 (-1.68,0.63) |
| Change 7-12yr (bpm/yr) | -1.89 (-2.08,-1.71) | 0.23 (0.00,0.47) | 0.13 (-0.12,0.37) | 0.00 (-0.27,0.28) |
| Change 12-16yr (bpm/yr) | -0.18 (-0.46,0.10) | -0.11 (-0.46,0.24) | -0.10 (-0.47,0.27) | 0.04 (-0.37,0.45) |
| Change 16-18yr (bpm/yr) | -4.09 (-4.69,-3.48) | -0.37 (-1.13,0.39) | -1.05 (-1.86,-0.24) | -1.20 (-2.10,-0.30) |
| Age 18yr (bpm) | 68.14 (67.28,69.00) | -0.76 (-1.85,0.33) | -2.74 (-3.91,-1.58) | -2.74 (-4.03,-1.45) |
| **Males** | | | | |
| Age 7yr (bpm) | 82.65 (81.91,83.39) | 0.07 (-0.87,1.01) | 0.01 (-0.99,1.02) | 0.09 (-1.07,1.24) |
| Change 7-12yr (bpm/yr) | -1.94 (-2.12,-1.75) | -0.02 (-0.25,0.21) | -0.02 (-0.26,0.23) | 0.10 (-0.18,0.38) |
| Change 12-16yr (bpm/yr) | -0.88 (-1.18,-0.58) | 0.25 (-0.12,0.63) | 0.17 (-0.22,0.55) | 0.34 (-0.09,0.77) |
| Change 16-18yr (bpm/yr) | -3.54 (-4.23,-2.85) | -0.58 (-1.44,0.29) | -0.62 (-1.51,0.27) | -1.33 (-2.31,-0.35) |
| Age 18yr (bpm) | 62.39 (61.37,63.41) | -0.16 (-1.43,1.12) | -0.64 (-1.96,0.67) | -0.73 (-2.17,0.71) |

DBP, diastolic blood pressure; SBP, systolic blood pressure; CI, confidence interval

[a]Units are presented in mmHg for SBP and DBP and bpm for pulse rate

**Table S13:** Mean trajectories of glucose and mean differences by maternal education, estimated from multilevel models

| | Mean trajectory (95% CI) in less than O-level maternal education (reference) | Mean difference in trajectory (95% CI) comparing with less than O-level maternal education | | |
|---|---|---|---|---|
| | | O-level maternal education | A-level maternal education | Degree maternal education |
| <u>**Glucose**</u> | | | | |
| **Females** | | | | |
| Age 7yr (mmol/l) | 4.13 (4.09,4.17) | 0.02 (-0.04,0.07) | -0.01 (-0.07,0.05) | -0.05 (-0.11,0.02) |
| Change 7-15yr (mmol/l/yr) | 0.13 (0.12,0.14) | -0.004 (-0.01,0.01) | 0.003 (-0.01,0.01) | 0.01 (-0.003,0.02) |
| Change 15-18yr (mmol/l/yr) | -0.09 (-0.12,-0.07) | -0.001 (-0.03,0.03) | -0.01 (-0.04,0.02) | -0.01 (-0.05,0.02) |
| Age 18yr (mmol/l) | 4.91 (4.86,4.96) | -0.01 (-0.07,0.05) | -0.02 (-0.08,0.05) | -0.01 (-0.08,0.06) |
| **Males** | | | | |
| Age 7yr (mmol/l) | 4.21 (4.17,4.25) | -0.03 (-0.08,0.02) | -0.004 (-0.06,0.05) | -0.04 (-0.10,0.03) |
| Change 7-15yr (mmol/l/yr) | 0.14 (0.13,0.15) | 0.004 (-0.01,0.01) | -0.0003 (-0.01,0.01) | 0.003 (-0.01,0.02) |
| Change 15-18yr (mmol/l/yr) | -0.07 (-0.09,-0.04) | -0.02 (-0.05,0.02) | -0.004 (-0.04,0.03) | -0.02 (-0.06,0.01) |
| Age 18yr (mmol/l) | 5.16 (5.10,5.21) | -0.05 (-0.11,0.02) | -0.02 (-0.08,0.05) | -0.09 (-0.16,-0.02) |

CI, confidence interval; HDL-c, high density lipoprotein cholesterol; mmol/l, millimole per litre; %/yr, percentage per year

**Table S14:** Mean trajectories of HDL-c and non-HDL-c and mean differences by maternal education, estimated from multilevel models

| | Mean trajectory (95% CI) in less than O-level maternal education (reference) | Mean difference in trajectory (95% CI) comparing with less than O-level maternal education | | |
|---|---|---|---|---|
| | | O-level maternal education | A-level maternal education | Degree maternal education |
| <u>**HDL-c**</u> | | | | |
| **Females** | | | | |
| Birth (mmol/l) | 0.54 (0.52,0.56) | -0.02 (-0.04,0.01) | 0.02 (-0.01,0.05) | 0.03 (-0.001,0.06) |
| Change 0-7yr (mmol/l/yr) | 0.13 (0.13,0.14) | 0.004 (-0.001,0.01) | -0.004 (-0.01,0.002) | -0.004 (-0.01,0.003) |
| Change 7-18yr (mmol/l/yr) | -0.02 (-0.02,-0.01) | 0.002 (-0.002,0.01) | 0.01 (0.002,0.01) | 0.01 (0.01,0.01) |
| Age 18yr (mmol/l) | 1.28 (1.25,1.30) | 0.03 (0.001,0.06) | 0.05 (0.02,0.08) | 0.11 (0.07,0.15) |
| **Males** | | | | |
| Birth (mmol/l) | 0.49 (0.47,0.51) | 0.02 (0.0003,0.05) | 0.02 (-0.002,0.05) | 0.01 (-0.02,0.04) |
| Change 0-7yr (mmol/l/yr) | 0.15 (0.15,0.16) | -0.005 (-0.01,-0.0003) | -0.01 (-0.01,-0.0003) | -0.01 (-0.01,0.001) |
| Change 7-18yr (mmol/l/yr) | -0.04 (-0.04,-0.03) | 0.001 (-0.003,0.004) | 0.002 (-0.002,0.01) | 0.003 (-0.001,0.01) |
| Age 18yr (mmol/l) | 1.15 (1.12,1.18) | -0.01 (-0.04,0.03) | 0.002 (-0.03,0.04) | 0.005 (-0.03,0.04) |
| <u>**Non-HDL-c**</u> | | | | |
| **Females** | | | | |
| Birth (mmol/l) | 1.28 (1.23,1.32) | 0.01 (-0.05,0.06) | 0.06 (-0.01,0.12) | 0.02 (-0.05,0.10) |
| Change 0-9yr (mmol/l/yr) | 0.21 (0.20,0.22) | -0.003 (-0.01,0.01) | -0.01 (-0.02,0.003) | -0.01 (-0.03,-0.002) |
| Change 9-18yr (mmol/l/yr) | -0.07 (-0.08,-0.06) | -0.006 (-0.01,0.01) | -0.01 (-0.01,0.004) | 0.002 (-0.01,0.01) |
| Age 18yr (mmol/l) | 2.55 (2.49,2.61) | -0.05 (-0.12,0.02) | -0.09 (-0.16,-0.01) | -0.09 (-0.18,-0.01) |
| **Males** | | | | |
| Birth (mmol/l) | 1.22 (1.18,1.26) | 0.002 (-0.05,0.06) | 0.02 (-0.05,0.08) | 0.01 (-0.07,0.09) |
| Change 0-9yr (mmol/l/yr) | 0.19 (0.19,0.20) | -0.002 (-0.01,0.01) | -0.005 (-0.01,0.01) | -0.003 (-0.01,0.01) |
| Change 9-18yr (mmol/l/yr) | -0.07 (-0.08,-0.06) | -0.01 (-0.01,0.004) | -0.002 (-0.01,0.01) | 0.001 (-0.01,0.01) |
| Age 18yr (mmol/l) | 2.32 (2.26,2.38) | -0.06 (-0.13,0.02) | -0.04 (-0.13,0.03) | -0.01 (-0.9,0.08) |

CI, confidence interval; HDL-c, high density lipoprotein cholesterol; mmol/l, millimole per litre

**Table S15:** Mean trajectories of triglycerides by maternal education, estimated from multilevel models

| | Mean trajectory (95% CI) in less than O-level maternal education (reference)[a] | Mean difference in trajectory (95% CI) comparing with less than O-level maternal education [b] | | |
|---|---|---|---|---|
| | | O-level maternal education | A-level maternal education | Degree maternal education |
| **Female** | | | | |
| Birth (mmol/l or %) | -0.70 (-0.73,-0.66) | 2.40 (-2.43,7.22) | 4.68 (-0.93,10.28) | 0.31 (-5.98,6.60) |
| Change 0-9yr (mmol/l/yr or %/yr) | 0.09 (0.08,0.10) | -0.45 (-1.17,0.27) | -0.56 (-1.36,0.24) | -0.23 (-1.16,0.69) |
| Change 9-18yr (mmol/l/yr or %/yr) | -0.04 (-0.05,-0.04) | 0.09 (-0.58,0.75) | -0.33 (-1.02,0.36) | -0.62 (-1.37,0.14) |
| Age 18yr (mmol/l or %) | -0.26 (-0.30,-0.23) | -0.91 (-5.36,3.54) | -3.37 (-7.91,1.17) | -7.11 (-11.86,-2.35) |
| **Male** | | | | |
| Birth (mmol/l or %) | -0.68 (-0.72,-0.65) | 1.31 (-3.37,6.00) | 0.14 (-5.03,5.31) | 3.53 (-3.19,10.26) |
| Change 0-9yr (mmol/l/yr or %/yr) | 0.08 (0.07,0.08) | -0.10 (-0.81,0.61) | 0.32 (-0.45,1.10) | 0.04 (-0.90,0.98) |
| Change 9-18yr (mmol/l/yr or %/yr) | -0.04 (-0.04,-0.03) | -0.23 (-0.91,0.45) | -0.52 (-1.21,0.18) | -0.31 (-1.08,0.46) |
| Age 18yr (mmol/l or %) | -0.32 (-0.36,-0.29) | -1.66 (-6.40,3.08) | -1.59 (-6.37,3.19) | 1.07 (-4.30,6.45) |

CI, confidence interval; mmol/l, millimole per litre; mmol/l/year; %/yr, percentage per year

[a]Triglyceride was transformed using the natural log. All predicted mean values (mmol/l) and rates of change per year (mmol/l/yr) are on the log scale

[b]The difference by maternal education is back transformed from the log scale for ease of interpretation and is interpreted as the percentage difference in the mean level comparing each category with less than O-level maternal education or percentage difference in change per year (%/yr) comparing each category with less than O-level maternal education
